# The causes and consequences of Alzheimer’s disease: phenome-wide evidence from Mendelian randomization

**DOI:** 10.1101/2019.12.18.19013847

**Authors:** Roxanna Korologou-Linden, Laura D Howe, Louise A C Millard, Yoav Ben-Shlomo, Dylan M Williams, George Davey Smith, Emma L Anderson, Evie Stergiakouli, Neil M Davies

## Abstract

**Importance:** Alzheimer’s disease is the leading cause of disability and healthy life years lost. However, to date, there are no proven causal and modifiable risk factors, or effective interventions.

**Objective:** We aimed to identify: a) factors modified by prodromal Alzheimer’s disease pathophysiology and b) causal risk factors for Alzheimer’s disease. We identified factors modified by Alzheimer’s disease using a phenome-wide association study (PheWAS) on the Alzheimer’s disease polygenic risk score (PRS) (p≤5×10^-8^), stratified by age tertiles. We used two-sample bidirectional Mendelian randomization (MR) to estimate the causal effects of identified risk factors and correlates on liability for Alzheimer’s disease.

**Design, setting, and participants:** 334,968 participants of the UK Biobank aged 39 to 72 years old (111,656 in each tertile) met our eligibility criteria.

**Exposures:** Standardized weighted PRS for Alzheimer’s disease at p≤5×10^-8^.

**Main outcomes and measures:** All available phenotypes in UK Biobank, including data on health and lifestyle, as well as samples from urine, blood and saliva, at the time of analysis.

**Results:** Genetic liability for Alzheimer’s disease was associated with red blood cell indices and cognitive measures at all ages. In the middle and older age tertiles, ages 53 and above, higher genetic liability for Alzheimer’s disease was adversely associated with medical history (e.g. atherosclerosis, use of cholesterol-lowering medications), physical measures (e.g. body fat measures), blood cell indices (e.g. red blood cell distribution width), cognition (e.g. fluid intelligence score) and lifestyle (e.g. self-reported moderate activity). In follow-up analyses using MR, there was only evidence that education, fluid intelligence score, hip circumference, forced vital capacity, and self-reported moderate physical activity were likely to be causal risk factors for Alzheimer’s disease.

**Conclusion and relevance:** Genetic liability for Alzheimer’s disease is associated with over 160 phenotypes, some as early as age 39 years. However, findings from MR analyses imply that most of these associations are likely to be a consequence of prodromal disease or selection, rather than a cause of the disease.

## INTRODUCTION

Alzheimer’s disease is a late-onset irreversible neurodegenerative disorder, constituting the majority of dementia cases.^1^ Despite major private and public investments in research, there are no effective treatments for preventing the disease.^2^ Many risk factors and biomarkers have been identified to associated with risk of Alzheimer’s disease.^3^ 99.6% of treatments developed to halt Alzheimer’s disease failed in phase I, II, or III trials.^4^ One explanation for these failures are that the identified risk factors and drug targets are a consequence of Alzheimer’s disease, rather than its underlying cause.

Genetic epidemiologic methods, such as Mendelian randomization (MR) can potentially provide more reliable insights into the causal mechanisms underlying the associations between risk factors and disease.^5^ To date, hypothesis-driven MR studies have found mixed evidence for a causal role of cardiovascular risk factors in the development of Alzheimer’s disease.^6-8^

Phenome-wide association studies (PheWAS) are a hypothesis-free method, similar to genome-wide association studies (GWAS), which estimate the associations between a genotype or polygenic risk score (PRS) and the phenome.^9^ PheWAS elucidate the phenotypic consequences of Alzheimer’s disease, and critically when in the life course these effects emerge. We estimated the associations of genetic liability for Alzheimer’s disease and the phenome by age (**Fig 1a**). We then tested whether the identified phenotypes were a cause or a consequence of Alzheimer’s disease using bidirectional MR (**Fig 1b**).

**Fig 1.**
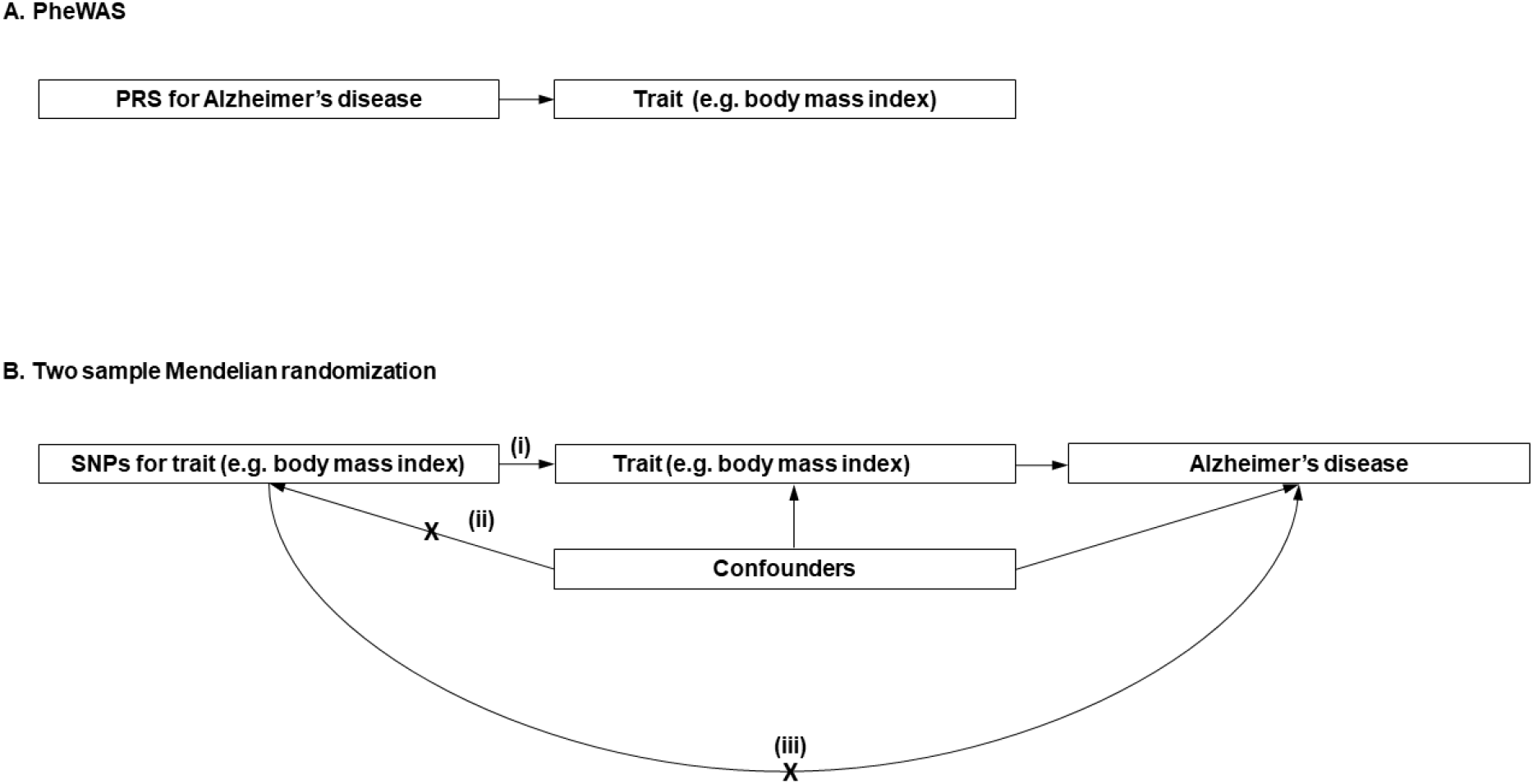
Diagram (A) describes our study design when conducting a phenome-wide association study (PheWAS) and diagram (B) describes our study design when using MR. In (A), the PRS for Alzheimer’s disease may either have a downstream causal effect on the trait (e.g. body mass index), or it may affect the trait through pathways other than through Alzheimer’s disease (i.e. pleiotropic effects). Diagram (B) describes our follow-up analysis using MR to establish causality and directionality of the observed associations. In (B), we test the hypothesis that the trait (e.g. body mass index) causally affects liability to Alzheimer’s disease, provided that the conditions (i), (ii), and (iii) are satisfied. The PRS for the trait of interest is a valid instrument, in that (i) the SNPs for a trait are strongly associated with the trait they proxy (relevance), (ii) there are no confounders of the SNPs-outcome relationship (independence), and (iii) the SNPs only affect the outcome via their effects on the trait of interest (exclusion restriction).

## METHODS

### Study design

Our analysis proceeded in two steps. First, we ran a PheWAS of the Alzheimer’s disease PRS and all available phenotypes in UK Biobank, stratifying the sample by age. Second, we followed-up all phenotypes associated with the PRS using two-sample MR. We outline the research questions answered by the PheWAS and the MR approach in **Fig 1**.

### Sample description

UK Biobank is a population-based study of 503,325 people recruited between 2006 and 2010 from across Great Britain.^10,11^ A full description of the study design, participants and quality control (QC) methods have been published previously.^11^ In total, a sample of 334,968 remained after QC (Supplementary Fig 1).

### Polygenic Risk Score

We constructed a standardized weighted PRS including single nucleotide polymorphisms (SNPs) associated with Alzheimer’s disease at p≤5×10^-8^ for UK Biobank participants, based on the summary statistics from a meta-analysis of the IGAP consortium,^12^ ADSP ^13^ and PGC ^14^ (24,087 cases and 55,058 controls) (details in Supplementary material). Our main analysis used the PRS including variants near the *APOE* gene (Chr 19: 44,400 kb-46,500 kb).^15^ The *APOE* region explains a large proportion of the variance in the polygenic risk score (R^2^=84%). The PRS was standardized by subtracting the mean and dividing by the standard deviation (SD) of the PRS.

### Main analysis

The full UK Biobank sample was divided into three subsamples (n=111,656 in each tertile). We performed PheWAS within each tertile. Age, sex and first 10 genetic principal components were included as covariates.

### Outcomes

The Biobank data showcase enables researchers to identify variables based on the field type (http://biobank.ctsu.ox.ac.uk/showcase/list.cgi). There were 2,655 fields of the following types: integer, continuous, categorical (single and multiple). We excluded 55 fields *a priori* including age and sex, and technical variables (e.g. assessment center) (**Supplementary Table 2**).

## STATISTICAL ANALYSES

### Phenome-wide association study

We estimated the association of an Alzheimer’s disease PRS with each phenotype in the three age strata using PHESANT (version 14). A description of PHESANT’s automated rule-based method is published elsewhere.^16^ We accounted for the multiple tests performed by generating adjusted p-values, controlling for a 5% false discovery rate. The threshold (≤0.05) was used as a heuristic to identify phenotypes to follow-up in the MR analysis and not as an indicator of significance.^17,18^ Categories for the ordered categorical variables are in Supplementary Table 3.

### Sensitivity analysis

We repeated the PheWAS for the entire sample without stratifying by age to maximize power to detect associations. Furthermore, to examine if any detected associations could be attributed to the variants in or near the *APOE* gene, we repeated the PheWAS on the entire sample excluding this region from the PRS.

### Follow-up using MR

We investigated whether the phenotypes identified in our PheWAS or previous reported risk factors ^3^ were a cause or consequence of Alzheimer’s disease using bidirectional two-sample MR (details in Supplementary material). For each risk factor identified by the PheWAS (in ages 62-72 years) and literature reviews, we identified SNPs that are strongly associated (p≤5×10^-8^) with each trait. SNPs in the *APOE* region ^19^ were removed from instruments proxying the exposures.

### Alzheimer’s disease GWAS

We used the same meta-analysis of the IGAP consortium,^12^ ADSP ^13^ and PGC as described above for the two-sample MR analyses.^14^

### Estimating the effect of risk factors on Alzheimer’s disease

We harmonized the exposure and outcome GWAS (details in Supplementary material). We estimated the effect of each exposure on Alzheimer’s disease using MR and the inverse-variance weighted (IVW) estimator; this estimator assumes no directional horizontal pleiotropy.^5^ We used the F-statistic as a measure of instrument strength.^20^ We present adjusted p-values for inverse variance weighted regression accounting for the number of results in the follow-up using the false discovery rate method.

### Assessing pleiotropy

We investigated whether the SNPs had pleiotropic effects on the outcome other than through the exposure using Egger regression.^21,22^ Egger regression allows for pleiotropic effects that are independent of the effect on the exposure of interest.^21,23,24^ We also report the I^2^G_x_ statistic, ^25^ an analogous measure to the F-statistic in inverse variance weighted regression.

### Assessing causal direction

We used Steiger filtering to investigate the direction of causation between Alzheimer’s disease and the phenotypes.^26^ Steiger filtering examines whether the SNPs for each of the phenotypes used in the two-sample MR explain more variance in the phenotypes than in Alzheimer’s disease (which should be true if the hypothesized direction from phenotype to Alzheimer’s disease is valid). We repeated MR analyses removing SNPs which explained more variance in the outcome than in the exposure.

### Data and code availability

The data in the current study were partly provided by the UK Biobank Study (www.ukbiobank.ac.uk), received under UK Biobank application no. 16729. Scripts are available on Github at: https://github.com/rskl92/AD_PHEWAS_UKBIOBANK.

## RESULTS

### Sample characteristics

The UK Biobank sample is 55% female (39 to 53 years, mean=47.2 years, SD=3.8 years) in tertile 1, 55% female (53 to 62 years, mean=58.03 years, SD=2.4 years) in tertile 2 and 49% female (62 to 72 years, mean=65.3 years, SD=2.7 years) in tertile 3. In the whole sample, the Alzheimer’s disease PRS was associated with a lower age at recruitment (β: −0.006 years; 95% CI: −0.01, −0.0002; p=0.007). The mean standardized PRS (95% CI) in each tertile was as follows: 0.006 (−0.0003, 0.01); and 0.001 (−0.01, 0.009) and −0.007 (−0.02, 0.002) (P for trend=0.01).

### Association of Alzheimer’s disease PRS and the phenome

Selected PheWAS hits are presented in graphs and full results are in Supplementary material. Results for continuous outcomes are in terms of a 1 SD change of inverse rank normal transformed outcome and log odds or odds for binary or categorical outcomes. A higher PRS for Alzheimer’s disease was associated with own diagnosis and family history of dementia, diagnoses of cardiovascular diseases, as well as a self-reported history of high cholesterol and pure hypercholesterolaemia. Furthermore, participants with a higher PRS had an increased risk of using cholesterol-lowering drugs, in addition to beta-blockers and aspirin in all age tertiles (Supplementary Fig 3). A higher PRS was associated with lower body mass index as well as various body fat measures, lower diastolic blood pressure and higher spherical power in the oldest participants (i.e. strength of lens needed to correct focus) (Fig. 2). Additionally, participants with higher PRS, on average, performed worse and took longer to complete cognitive tests in all ages examined (39 to 72 years) (Fig 3). Participants with a higher PRS also had a higher weighted-mean mode of anisotropy in the left inferior fronto-occipital fasciculus for participants aged 53 to 72 years (Fig 3). There was evidence of association between higher PRS and blood cell composition markers, where effects for most of these increased with age (Fig 4). On average, the parents of participants with a higher PRS for Alzheimer’s disease died at a younger age (Supplementary Fig 4). There was strong evidence that a higher PRS was associated with healthier dietary choices (Supplementary Fig 4) and lifestyles (e.g. frequent exercise) in two oldest tertiles (ages 53-72 years) (Supplementary Fig 5). For previously implicated factors in Alzheimer’s disease, a higher PRS was associated with higher systolic blood pressure only in participants aged 39-53 years and higher pulse pressure for all age ranges. There was some evidence of an association between the PRS and lower number of pack years of smoking for the oldest participants (Supplementary Fig 5).

**Fig 2.**
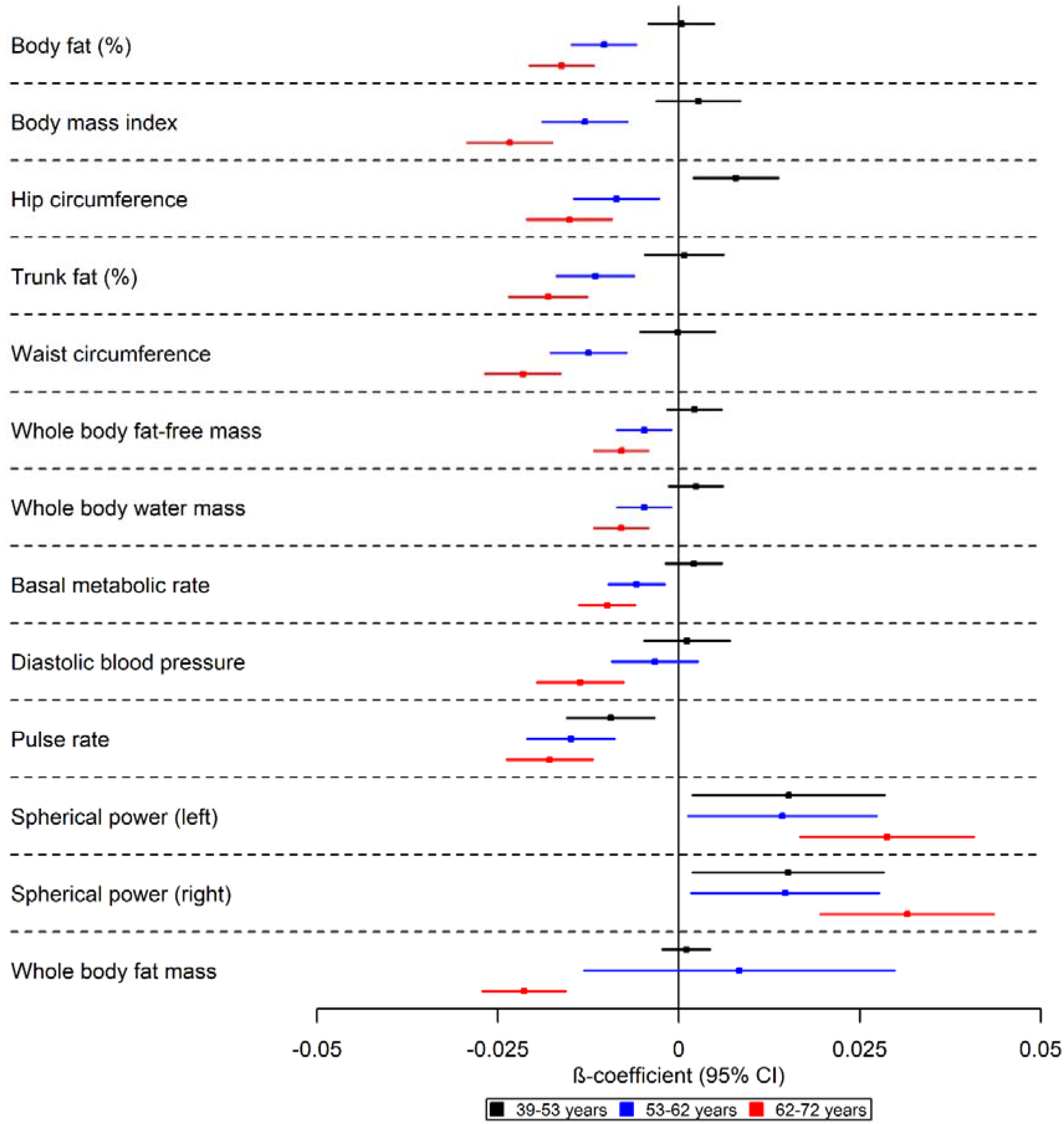
Forest plot showing the effect estimates for the association between the polygenic score for Alzheimer’s disease (including the apolipoprotein E region) and physical measures. Legends in the right of each graph indicate age tertiles. Effect estimates are shown by box markers and confidence bands represent 95% confidence intervals. There is evidence that the PRS for Alzheimer’s disease is related to physical measures in older, but not younger participants. This suggests that Alzheimer’s disease causes these changes rather than vice versa.

**Fig 3.**
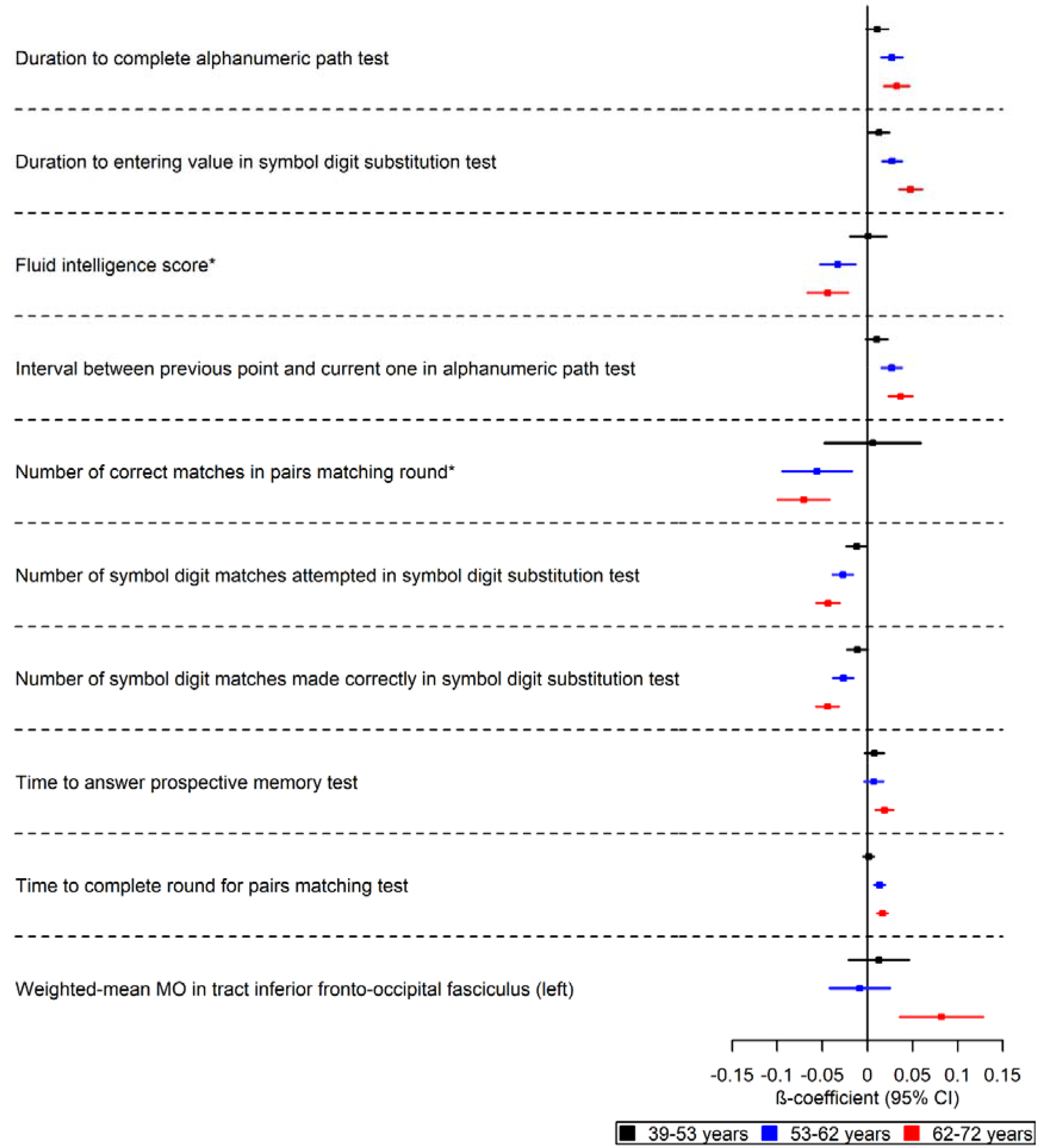
Forest plot showing the effect estimates for the association between the polygenic score for Alzheimer’s disease (including the apolipoprotein E region), cognitive, and brain-related measures. Legends in the right of each graph indicate age tertiles. Effect estimates are shown by box markers and confidence bands represent 95% confidence intervals. *Effect estimates were derived from ordered logistic models and effect estimates are on log odds scale. We found evidence that the PRS for Alzheimer’s disease is related to some cognitive measures in all age ranges examined. This may suggest a bidirectional relationship between cognitive measures and Alzheimer’s disease.

**Fig 4.**
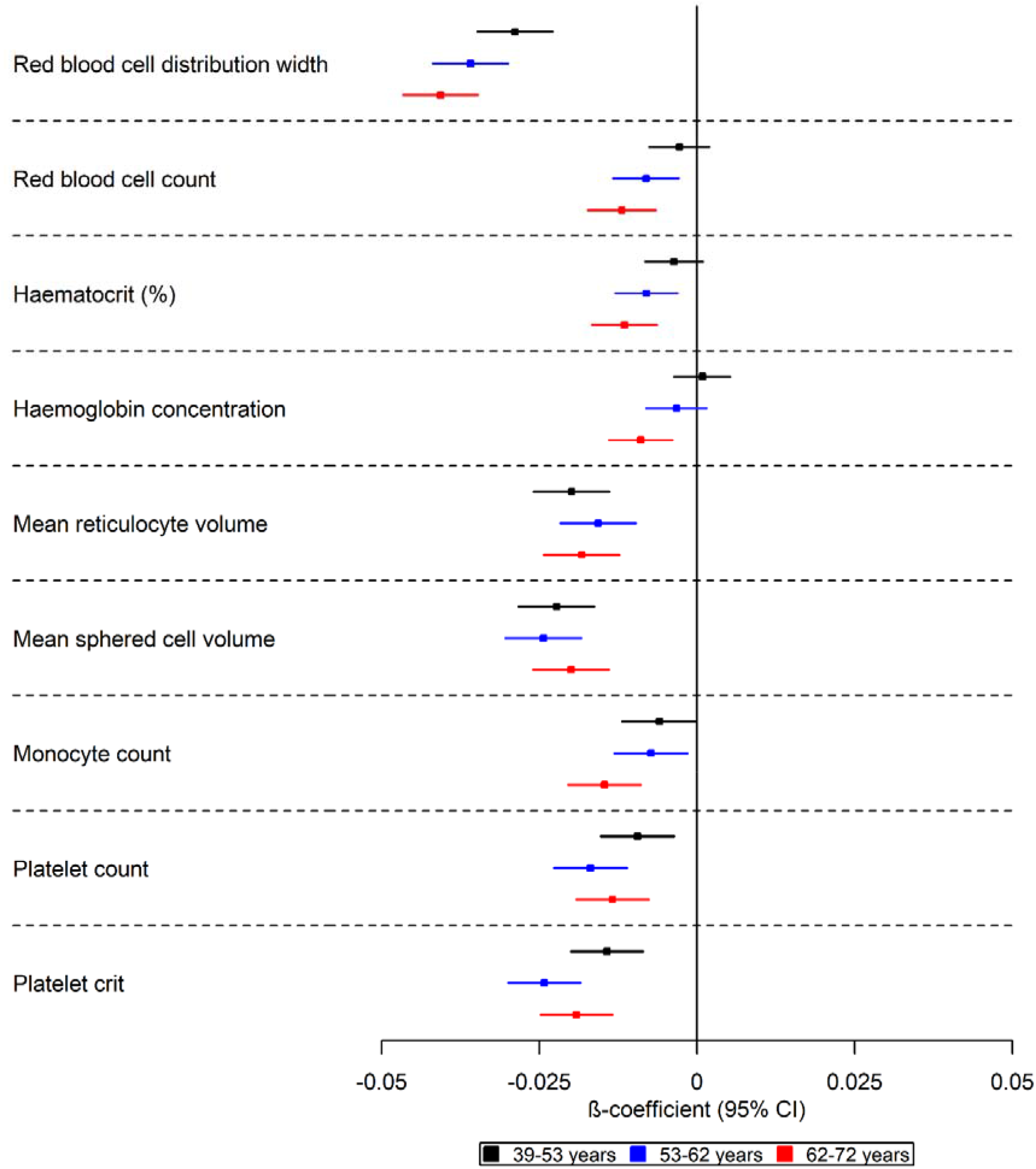
Forest plot showing the effect estimates for the association between the polygenic score for Alzheimer’s disease (including the apolipoprotein E region) and biological measures. Legends in the right of each graph indicate age tertiles. Effect estimates are shown by box markers and confidence bands represent 95% confidence intervals. There is an age-dependent increase in the effect of the polygenic risk score on blood-based measures. This may indicate that blood-based markers may be causal in the development of Alzheimer’s disease.

### Sensitivity analysis

We repeated the analysis estimating the associations of the PRS and the phenotypes for the entire sample. We detected associations with more phenotypes (Supplementary Figs 6-9), a higher PRS was associated with metabolic dysfunction phenotypes such as diabetes and obesity (Supplementary Fig 6); a higher volume of grey matter in right and left intracalcarine and supracalcarine cortices (Supplementary Fig 8); and additional blood-based biomarkers (Supplementary Fig 8).

When we repeated the analysis removing SNPs tagging the *APOE* region from the PRS using the whole sample, we could not replicate most of the hits detected in the oldest tertile. The *non-APOE* PRS was associated with higher odds of own and family diagnosis of Alzheimer’s disease (Supplementary Fig 10), in addition to lower odds of family history of chronic bronchitis/emphysema (Supplementary Fig 11). There was evidence that the non-APOE PRS was associated with worse performance in cognitive tests (Supplementary Fig 12).

### Two-sample MR of UK Biobank phenotypes on Alzheimer’s disease

We found evidence that a one SD higher genetically predicted hip circumference decreased the risk of Alzheimer’s disease (OR: 0.75; 95% CI:0.61,0.90) and that a one SD higher genetically predicted forced vital capacity resulted in 22% lower odds of risk for Alzheimer’s disease (OR: 0.78; 95% CI:0.67,0.90) (Supplementary table 12). A one SD higher genetically predicted fluid intelligence score reduced the odds of Alzheimer’s disease by 27% (OR: 0.73; 95% CI:0.59,0.90) (Supplementary table 13). We observed that a higher genetic liability of doing more moderate physical activity (at least 10 minutes), but not self-reported vigorous activity, increased the odds of developing Alzheimer’s disease (OR: 2.29; 95% CI:1.32, 3.98 and OR: 1.02; 95% CI: 0.33,3.18, respectively) (Supplementary table 16). For previously implicated risk factors for Alzheimer’s disease, we found a higher genetic liability for having a college degree and A level qualifications reduced risk of Alzheimer’s disease.

### Assessing pleiotropy

Due to the large sample size of the exposures, the instrument strength of all SNPs was relatively high (F>30). However, the SNPs used for each body measurement implied highly heterogeneous effects on risk of Alzheimer’s disease (all heterogeneity Q statistic P<3.37×10^-5^). This heterogeneity may be due to horizontal pleiotropy (Supplementary Figs 14-18). The causal effect estimates for forced vital capacity and fluid intelligence score were also heterogenous (Supplementary tables 12,13 and Figs 19, 20).

### Assessing causal direction

We assessed the causal direction using Steiger filtering. We found little evidence that SNPs explained more variance in the outcome than the exposure for most results reported in the two-sample MR section. However, the effect estimates for MR analyses retaining only the SNPs with the true hypothesized causal direction attenuated for body fat percentage, whole body fat and fat-free mass **(Supplementary Materials)**.

### Discussion

To our knowledge, this is the first study to conduct a hypothesis-free phenome-wide scan to investigate both how, and at what age, the Alzheimer’s disease PRS affects the phenome. The effects of a higher genetic liability for Alzheimer’s disease are stronger in participants of ages 62 to 72 years although the direction of effect is largely similar across age groups. We investigated whether the effects observed are causes or consequences of the disease process using two-sample bidirectional MR. We found evidence that a minority of traits are likely to casually affect liability to Alzheimer’s **(Fig 5)**.

**Fig 5.**
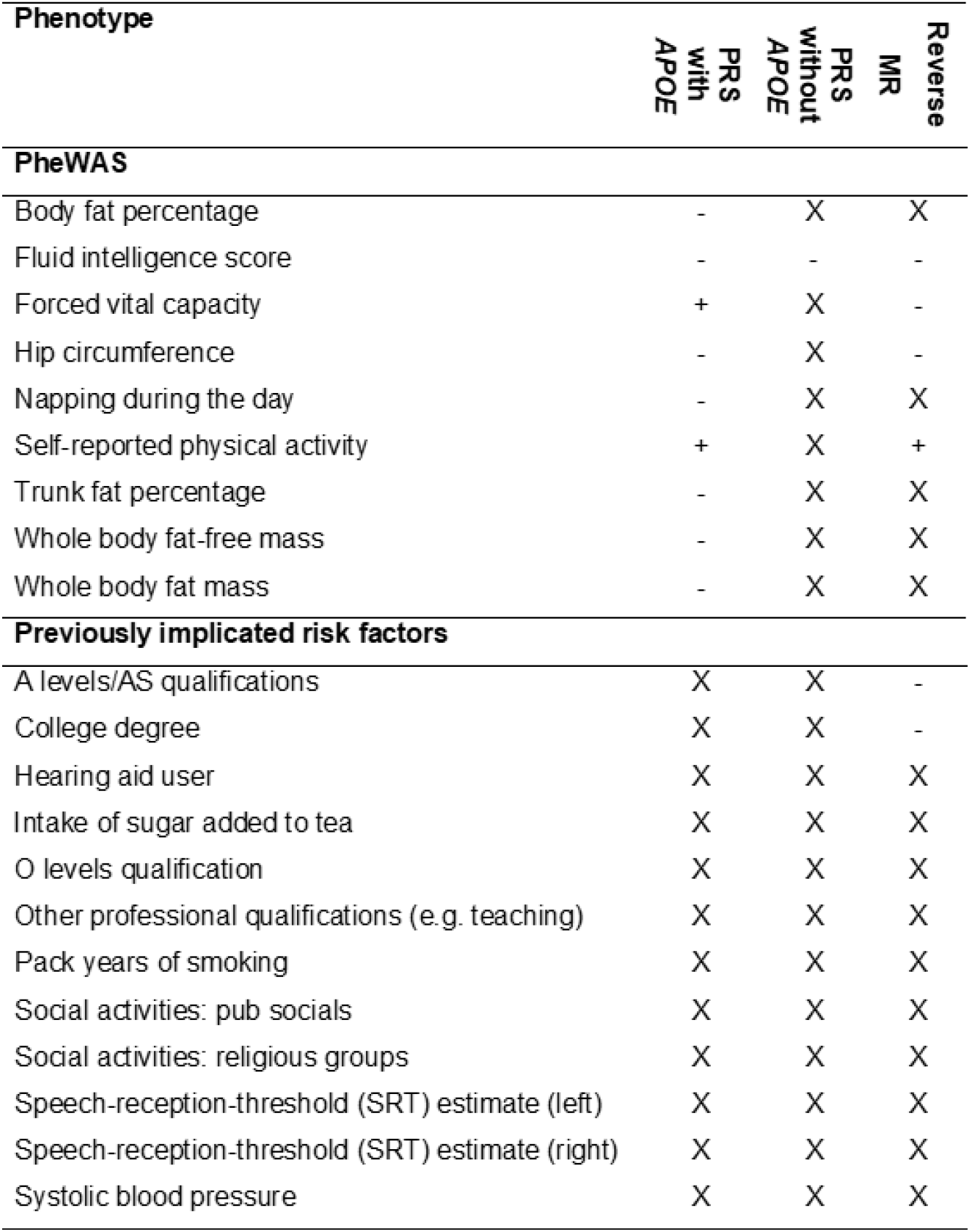
Association of Alzheimer’s disease PRS with phenome, and estimated effect of each phenotype using MR. + and – indicate the direction of the coefficient for phenotypes associated with Alzheimer’s disease using two-sample MR. X represents associations which were consistent with the null.

The PheWAS using all Alzheimer’s disease SNPs (including those in the *APOE* gene) suggested that increased genetic liability for Alzheimer’s disease affected a diverse array of phenotypes such as medical history, brain-related phenotypes, physical measures, lifestyle, and blood-based measures. However, these effects appear to be largely driven by variation in the *APOE* gene, as our sensitivity analysis excluding the *APOE* region replicated only effects for family history of Alzheimer’s disease and some cognitive measures. These results are consistent with observational studies ^27-33^ and studies in APOE-deficient mice, ^34-37^ which demonstrate the multifunctional role of *APOE* on longevity-related phenotypes such as changes in lipoprotein profiles, ^34,38-40^ neurological disorders,^41^ type II diabetes,^35^ altered immune response,^36^ and increased markers of oxidative stress.^37^ Therefore, this strongly suggests that the effects of the Alzheimer’s PRS on the phenome (e.g. atherosclerotic heart disease) are likely to be due to biological pathways related to *APOE*.

Previous observational studies have reported conflicting evidence on the association of cardiovascular risk factors including hypertension with Alzheimer’s disease. The results have depended on the age at which these risk factors were measured.^42-44^ Similarly, in our study, a higher PRS for Alzheimer’s disease was associated with lower body mass index and body fat in older participants as well as lower diastolic blood pressure. In agreement with some previous MR studies,^6,7,45,46^ we found little evidence that body mass index and blood pressure causally affect risk of developing Alzheimer’s disease. Hence, the association observed in the PheWAS between the PRS, lower body fat measures and diastolic blood pressure is likely to reflect the prodromal disease process. We found evidence that a higher self-reported number of days of moderate physical activity increased the odds of Alzheimer’s disease. A MR study ^47^ also found evidence that a higher moderate vigorous physical activity was associated with a higher risk of Alzheimer’s disease and increased cerebrospinal fluid Aβ_42_ levels (indicative of higher cerebral amyloid load ^48-50^).

We found genetic risk for Alzheimer’s disease to be associated with several phenotypes involving inflammatory pathways such as self-reported wheeze/whistling, monocyte count, and blood-based measures. This is in agreement with previous evidence of genetic correlations between the Alzheimer’s disease and asthma ^51^ and longitudinal studies ^52,53^. In our study, red blood cell indices show the earliest evidence of association with the genetic risk of Alzheimer’s disease, but we found little evidence that these measures caused Alzheimer’s disease using MR indicating that cell composition changes may be an early consequence of Alzheimer’s disease pathophysiology. Previous studies found that genetic variants associated with red blood cell distribution width are linked to autoimmune disease and Alzheimer’s disease.^54,55^

We found evidence that a PRS for Alzheimer’s disease was associated with a lower fluid intelligence score, as previously reported ^56^ but not educational attainment. Although previous MR studies have suggested that higher educational attainment reduces liability for Alzheimer’s disease,^6,57,58^ a multivariable MR study found little evidence that educational attainment directly increased risk of Alzheimer’s disease over and above the underlying effects of intelligence.^59^

We identified suggestive evidence of a bidirectional relationship between sleep and Alzheimer’s disease. Self-reported daytime napping causally reduced risk of Alzheimer’s disease and a higher genetic risk for Alzheimer’s resulted in changed sleeping patterns such as lower sleep duration, higher frequency of sleeplessness/insomnia and lower frequency of daytime napping. Observational studies have shown that increased sleep fragmentation is associated with cognitive impairment and dementia ^60-62^ but the directionality of this relationship is unclear.^63^

### Strengths and limitations

The large sample of UK Biobank provided unparalleled statistical power to investigate the phenotypic manifestation of a higher genetic liability for Alzheimer’s disease, by age group. Furthermore, the systematic approach of searching for effects using PheWAS reduces bias associated with hypothesis-driven investigations.

The Alzheimer’s disease PRSs may have horizontal pleiotropic effects on different traits and disorders. However, here we are interested in all of the downstream consequences of increased genetic liability to Alzheimer’s disease including pleiotropic mechanism. Our results could be explained by collider bias, which may have been introduced into our study through selection of the study sample. The UK Biobank includes a highly selected, healthier sample of the UK population.^64^ Compared to the general population, participants are less likely to be obese, to smoke, to drink alcohol on a daily basis, and had fewer self-reported medical conditions.^65^ Selection bias may occur if those with a lower genetic liability to Alzheimer’s disease and a specific trait (e.g. higher education) are more likely to participate in the study. This could induce an association between genetic risk for Alzheimer’s disease and the traits in our study.^66^ Furthermore, if both the PRS for Alzheimer’s disease and the examined traits associate with survival, sampling only living people can induce spurious associations that do not exist in the general population.^67,68^ Such bias may have affected our findings for body mass, as individuals with higher body mass index and those with higher values of the Alzheimer’s PRS are less likely to survive and participate in UK Biobank. The PRS for Alzheimer’s disease in our analysis was associated with lower age at recruitment, suggesting that older people with higher values of the score are less likely to participate.

### Conclusion

In this phenome-wide association study, we identified that a higher genetic liability for Alzheimer’s disease is associated with 165 phenotypes of 15,403 UK Biobank phenotypes. Mendelian randomization analysis follow-up showed evidence that only six of these factors were implicated in the etiology of Alzheimer’s disease. We found little evidence that the remaining phenotypes examined are likely to modify the disease process, but rather the association with the Alzheimer’s disease PRS is likely to be due to reverse causation or selection bias. Further research should exploit the full array of potential relationships between the genetic variants implicated in Alzheimer’s disease, intermediate phenotypes, and clinical phenotypes by using other omics and phenotypic data to identify potential biological pathways changing the risk of Alzheimer’s disease.

## Question

How does higher genetic liability for Alzheimer’s disease affect the phenome across the life course and do any phenotypes causally affect incidence of disease?

## Findings

In this population-based cohort study of 334,968 participants, higher genetic risk for Alzheimer’s disease was associated with medical history (e.g. higher odds of diagnosis of atherosclerotic heart disease), cognitive (e.g. lower fluid intelligence score), physical (e.g. lower forced vital capacity) and blood-based measures (e.g. lower haematocrit) as early as 39 years of age.

## Meaning

Most of the identified phenotypes are likely to be symptoms of prodromal Alzheimer’s disease, rather than causal risk factors.

## Acknowledgments

This research has been conducted using the UK Biobank Resource under Application Number 16729. The MRC IEU UK Biobank GWAS pipeline was developed by B.Elsworth, R.Mitchell, C.Raistrick, L.Paternoster, G.Hemani, T.Gaunt (doi: 10.5523/bris.pnoat8cxo0u52p6ynfaekeigi).We acknowledge the members of the Psychiatric Genomics Consortium (PGC). The Alzheimer’s Disease Sequencing Project (ADSP) comprises 2 Alzheimer’s Disease (AD) genetics consortia and 3 National Human Genome Research Institute (NHGRI)-funded Large Scale Sequencing and Analysis Centers (LSAC). The 2 AD genetics consortia are the Alzheimer’s Disease Genetics Consortium (ADGC) funded by the NIA (U01 AG032984), and the Cohorts for Heart and Aging Research in Genomic Epidemiology (CHARGE) funded by the NIA (R01 AG033193), the National Heart, Lung, and Blood Institute (NHLBI), other NIH institutes, and other foreign governmental and nongovernmental organizations. The Discovery Phase analysis of sequence data is supported through UF1AG047133 (to G. Schellenberg, L.A. Farrer, M.A. Pericak-Vance, R. Mayeux, and J.L. Haines); U01AG049505 to S. Seshadri; U01AG049506 to E. Boerwinkle; U01AG049507 to E. Wijsman; and U01AG049508 to A. Goate. Data generation and harmonization in the Follow-up Phases is supported by U54AG052427 (to G. Schellenberg and Wang). The ADGC cohorts include Adult Changes in Thought (ACT), the Alzheimer’s Disease Centers (ADC), the Chicago Health and Aging Project (CHAP), the Memory and Aging Project (MAP), Mayo Clinic (MAYO), Mayo Parkinson’s Disease controls, the University of Miami, the Multi-Institutional Research in Alzheimer’s Genetic Epidemiology Study (MIRAGE), the National Cell Repository for Alzheimer’s Disease (NCRAD), the National Institute on Aging Late Onset Alzheimer’s Disease Family Study (NIA-LOAD), the Religious Orders Study (ROS), the Texas Alzheimer’s Research and Care Consortium (TARC), Vanderbilt University/Case Western Reserve University (VAN/CWRU), the Washington Heights-Inwood Columbia Aging Project (WHICAP) and the Washington University Sequencing Project (WUSP), the Columbia University Hispanic-Estudio Familiar de Influencia Genetica de Alzheimer (EFIGA), the University of Toronto (UT), and Genetic Differences (GD). The CHARGE cohorts with funding provided by 5RC2HL102419 and HL105756, include the following: the Atherosclerosis Risk in Communities (ARIC) Study which is conducted as a collaborative study supported by NHLBI contracts (HHSN268201100005C, HHSN268201100006C, HHSN268201100007C, HHSN268201100008C, HHSN268201100009C, HHSN268201100010C, HHSN268201100011C, and HHSN268201100012C), the Austrian Stroke Prevention Study (ASPS), the Cardiovascular Health Study (CHS), the Erasmus Rucphen Family Study (ERF), the Framingham Heart Study (FHS), and the Rotterdam Study (RS). The 3 LSACs are the Human Genome Sequencing Center at the Baylor College of Medicine (U54 HG003273), the Broad Institute Genome Center (U54HG003067), and the Washington University Genome Institute (U54HG003079). Biological samples and associated phenotypic data used in primary data analyses were stored at Study Investigators institutions and at the National Cell Repository for Alzheimer’s Disease (NCRAD, U24AG021886) at Indiana University funded by the NIA. Associated Phenotypic Data used in primary and secondary data analyses were provided by Study Investigators, the NIA-funded Alzheimer’s Disease Centers (ADCs), and the National Alzheimer’s Coordinating Center (NACC, U01AG016976) and the National Institute on Aging Genetics of Alzheimer’s Disease Data Storage Site (NIAGADS, U24AG041689) at the University of Pennsylvania, funded by the NIA and at the Database for Genotypes and Phenotypes (dbGaP) funded by the NIH. This research was supported in part by the Intramural Research Program of the NIH and the National Library of Medicine. Contributors to the Genetic Analysis Data included Study Investigators on projects that were individually funded by the NIA and other NIH institutes, and by private U.S. organizations, or foreign governmental or nongovernmental organizations. We also thank the International Genomics of Alzheimer’s Project (IGAP) for providing summary results data for these analyses. The investigators within IGAP contributed to the design and implementation of IGAP and/or provided data but did not participate in analysis or writing of this report. IGAP was made possible by the generous participation of the control subjects, the patients, and their families. The i-Select chips was funded by the French National Foundation on Alzheimer’s disease and related disorders. EADI was supported by the LABEX (laboratory of excellence program investment for the future) DISTALZ grant, Inserm, Institut Pasteur de Lille, Université de Lille 2 and the Lille University Hospital. GERAD was supported by the Medical Research Council (Grant n° 503480), Alzheimer’s Research UK (Grant n° 503176), the Wellcome Trust (Grant n° 082604/2/07/Z) and German Federal Ministry of Education and Research (BMBF): Competence Network Dementia (CND) grant n° 01GI0102, 01GI0711, 01GI0420. CHARGE was partly supported by the NIH/NIA grant R01 AG033193 and the NIA AG081220 and AGES contract N01-AG-12100, the NHLBI grant R01 HL105756, the Icelandic Heart Association, and the Erasmus Medical Center and Erasmus University. ADGC was supported by the NIH/NIA grants: U01 AG032984, U24 AG021886, U01 AG016976, and the Alzheimer’s Association grant ADGC-10-196728.

